# The Role of Distress-related Metabolic Dysfunction in Ovarian Cancer Development: a pooled case-control study

**DOI:** 10.64898/2026.08.27.26361473

**Authors:** Nan Lin, Raji Balasubramanian, Giulia Menichetti, A. Heather Eliassen, Britton Trabert, Julian Avila-Pacheco, Mary K. Townsend, Kathryn L. Terry, Clary B. Clish, Shelley S. Tworoger, Oana A. Zeleznik

## Abstract

**Background:** Evidence suggests chronic distress influences ovarian cancer (OC) etiology and metabolomic profiles. Here, we evaluated the association of a metabolite-based distress score (MDS) and OC risk.

**Methods:** We included two matched case-control studies nested within the Nurses’ Health Studies (N=584) and the Prostate, Lung, Colorectal, and Ovarian Cancer Screening Trial (N=348). Metabolites were measured 3-27 years before diagnosis using liquid-chromatography tandem mass spectrometry. We examined the association of quintiles of MDS and 19 constituent metabolites with OC risk using unconditional logistic regression and stratified by tumor histotype, menopausal status, and age at diagnosis.

**Results:** We observed women in the highest versus lowest quintile of MDS had an increased OC risk (OR=1.62,95%CI=1.03-2.54,p_trend_=0.07), and type 2 tumors (OR=1.71,95%CI=1.03-2.83,p_trend_=0.11). Associations were suggestively stronger for premenopausal and <69-year-old women, and driven by pseudouridine, and N2,N2- dimethylguanosine.

**Conclusion:** Our findings suggest chronic distress-associated metabolic dysregulation may represent a novel OC risk factor, especially among younger women.

## Introduction

Ovarian cancer is the sixth leading cause of cancer-related death among women in the United States and the seventh most fatal cancer globally.^1^ Identifying women at high risk remains a challenge, necessitating the identification of novel risk factors to improve risk prediction models.^2,3^

Emerging evidence suggests that chronic stress and psychological distress may contribute to ovarian cancer etiology, increasing risk. Distress is associated with prolonged activation of the sympathetic nervous system, which can promote ovarian tumor growth and foster a pro-inflammatory and immunosuppressed environment.^3–8^ Prior studies have demonstrated that distress conditions, such as depression and post-traumatic stress disorder, are linked to increased ovarian cancer risk, independent of co-occurring behavioral and health risk factors.^9–15^

Advancements in metabolomics now allow for precise measurement of small-molecule metabolites. Recent findings indicate that distress is associated with metabolic alterations and dysregulation.^16–18^ A previous study developed a metabolite-based distress score (MDS) composed of 20 metabolites associated with depression and anxiety,^18^ some of which have also been implicated in ovarian cancer risk.^19,20^ However, the relationship of the MDS and its constituent metabolites with ovarian cancer risk has not been comprehensively evaluated. Thus, we hypothesized that chronic distress-induced metabolic dysregulation increased the ovarian cancer risk, and leveraged metabolomic data collected 3 to 27 years prior to diagnosis in two nested case-control studies to assess the association of the MDS and its individual components with ovarian cancer risk.

## Methods

### Study design

Our analysis included two United States-based case-control studies, nested within Nurses Health Studies (NHS, NHSII)^21,22^ and Prostate, Lung, Colorectal and Ovarian Cancer Screening Trial (PLCO).^2^ NHS, established in 1976, enrolled 121,700 female registered nurses aged 30-55 years old, and NHSII, established in 1989, enrolled 116,429 women aged 25-42 years old. NHS/NHSII participants completed a baseline questionnaire and were followed by biennial questionnaires which collected updated information on their health conditions, lifestyle, reproductive factors and disease diagnoses. Heparin blood samples and accompanying short questionnaires were collected from 32,826 NHS participants (aged 43-69 years) between 1989 and 1990 and from 29,611 NHSII participants (aged 32-54 years) between 1996 and 1999.^22,23^ Part of the data from NHS and NHSII was published previously.^19,20^ PLCO, established in 1993, consisted of 155,000 women aged 55 to 74 years old without previous diagnosis of lung, colorectal, or ovarian cancer who were randomly assigned to either an intervention or usual care group. They completed a self-administered baseline questionnaire, including demographics, general risk factors, and medical histories.^2^ Participants in the ovarian intervention arm provided heparin blood samples at the baseline screening visit.^24^ NHS/NHSII study protocol was approved by the institutional review boards of the Brigham and Women’s Hospital and Harvard T.H. Chan School of Public Health, and those of participating registries as required. The protocol for this research was approved by the institutional review board of Mass General Brigham. The PLCO trial was approved by institutional review boards at the National Cancer Institute and PLCO study centers.

### Participants

For NHS/NHSII, cases of invasive and borderline epithelial ovarian cancer were matched to one living control with at least one intact ovary at time of blood collection on: cohort (NHS, NHSII); menopausal status and hormone therapy use at blood draw (premenopausal, postmenopausal/hormone therapy use, postmenopausal/no hormone therapy use, missing/unknown); menopausal status at diagnosis (premenopausal, postmenopausal, unknown); age at blood draw (±1year), date of blood collection (±1month); time of day of blood draw (±2hours); and fasting status (>8hours or ≤8hours). For PLCO, cases were matched to one living control with no history of oophorectomy at time of diagnosis on: age at blood collection (55-59, 60-64, 65-69, 70+years), race (white, black, other), study center, and time (a.m., p.m.) and date (3-month categories) of blood collection. All PLCO participants were postmenopausal at time of sample collection. In this study, ovarian cancer was diagnosed at least three years after the blood collection to reduce concerns about reverse causation (i.e., underlying invasive disease altering metabolomic profiles). All participants provided written informed consent.

### Procedure

We measured two batches of plasma metabolites at the Broad Institute (Cambridge, MA) using liquid chromatography-tandem mass spectroscopy (LC-MS) methods. All MDS metabolites included had coefficients of variation <25%, and passed delayed processing pilot study.^25^ Details of metabolite assay, batch-pooling were provided in the **Supplementary Materials** and **Supplementary Figure 1**.

### Outcomes

In NHS/NHSII, incident ovarian cancer cases were identified by self-report on questionnaires, report by family members, or linkage to registries (e.g., state cancer registries, National Death Index). Medical records or cancer registry data were reviewed by a gynecologic pathologist to classify cancer morphology, histology, grade, and stage. In PLCO, incident ovarian cancer cases were ascertained by annual questionnaires mailed to participants or linkage with population-based cancer registries and the National Death Index. Cases’ medical records were obtained by PLCO screening centers and used to extract ovarian tumor stage, histology, and grade. Date of death was obtained from the National Death Index for all studies and included a medical record review for cause of death.

### Statistical analysis

The statistical analysis pipeline is presented in **Figure 1** and described below. Details on the calculation of the **MDS** are provided in the **Supplementary Materials** and **Supplementary Figure 2**. The development and validation of the **fasting score** are described in the **Supplementary Materials**, **Supplementary Table 1**, and Supplementary Table 2.

**Figure 1.**
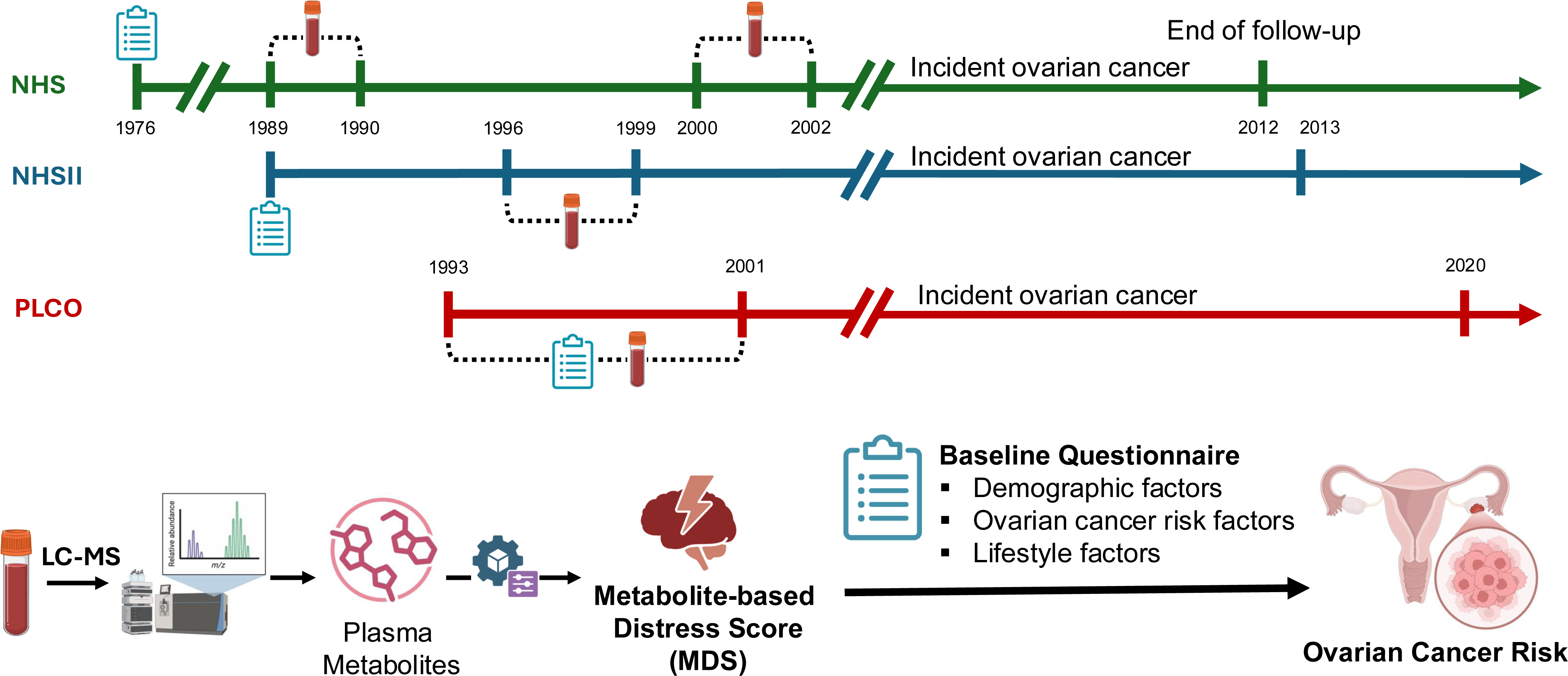
Flowchart of Study Analysis. Two matched case-control studies nested within the Nurses’ Health Studies (N=584) and the Prostate, Lung, Colorectal, and Ovarian Cancer Screening Trial (N=348). Metabolites were measured 3-27 years before diagnosis using liquid-chromatography tandem mass spectrometry. Detailed demographic, lifestyle, and dietary data were collected via questionnaires. The metabolite-based distress score (MDS) was developed as a weighted combination of 19 metabolites measured in plasma that is associated with chronic stress. We evaluated the association of ovarian cancer risk and quintiles of MDS and its 19 constituent metabolites using unconditional logistic regression adjusting for matching factors, ovarian cancer risk factors, and lifestyle factors.

Unconditional logistic regression was used to calculate odds ratios (OR) and 95% confidence intervals (95%) of ovarian cancer risk across quintiles of MDS (based on the distribution among controls), using the lowest quintile as reference group. We used median values across quintiles to calculate linear trend p-values. We considered three models. Model 1 included adjustment for the matching factors: age at blood collection (+/- 2years), study center, fasting status (yes, no), and menopausal status (premenopausal, postmenopausal, unknown). Model 2 included further adjustment for ovarian cancer risk factors: race and ethnicity (white, other), parity (none, 1, 2, 3, 4+ children), oral contraceptive use duration (never, <1year, 2-5years, 5+years), tubal ligation (yes, no), and family history of breast or ovarian cancer (yes, no). Model 3 additionally included adjustments for lifestyle factors and health behaviors that may be associated with distress or alterations in metabolite levels: smoking (former/current smoker, never), alcohol drinking (yes, no) body mass index (BMI, continuous), physical activity (<1, 1-2, 2-3, 3-4, 4+hours/week), and the aHEI (continuous). We also investigated the association of individual MDS metabolites (in quintiles based on control distributions) and ovarian cancer risk utilizing unconditional logistic regression and the same three statistical models presented above. Covariate assessment is described in **Supplementary Materials**.

We used polytomous logistic regression (PLR)^26^ to simultaneously estimate ORs and 95% CIs for ovarian cancer risk across ovarian cancer histotype (controls: n=466, type 1: low-grade serous, endometrioid, clear cell, mucinous; n=101 cases; type 2: high-grade serous, poorly differentiated, transitional/Brenner; n=290 cases) and time between MDS measurement and diagnosis for ovarian cancer cases with 3-12 years (234 cases) vs >12 years (232 cases). All PLR models were adjusted for the same covariates as the main logistic regression models. To evaluate heterogeneity of MDS quintiles in associations across tumor subtypes and by time between MDS measurement and diagnosis, we used likelihood ratio tests comparing a null model in which the associations for each MDS quintile were constrained to be equal across case groups, and an alternative model in which the MDS quintile associations were allowed to vary across groups based on the trend test.

Using the same models as in the main analysis, we conducted stratified analyses by known menopausal status at blood draw (premenopausal:99; postmenopausal:328 case-control sets), age at diagnosis/reference date for controls (<69years:204; ≥69 years:262 case-control sets), and fasting status with strata of fasting for at least eight hours (293cases/327controls) and not fasting (173cases/139controls). Wald tests were used to calculate heterogeneity in stratified analyses for each quintile (except the reference group) and across the linear trend. The analyses were conducted using SAS 9.4 and R version 4.2.0. A p-value of <0.05 was used to assess statistical significance.

## Results

A total of 584 participants from NHS/NHSII (292 case-control sets) and 348 participants from PLCO (174 case-control sets) were included in the analysis (**Table 1**). The median time between blood draw and ovarian cancer diagnosis was 11.8 years for NHS/NHSII and 8.3 years for PLCO. All PLCO participants were postmenopausal, whereas in NHS/NHSII, 53% were postmenopausal, 34% premenopausal, and 13% had unknown status at blood draw. Among NHS/NHSII cases, 80 were diagnosed with type 1 and 193 with type 2 ovarian cancer; other cases had unknown histotype. Among PLCO cases, 21 were diagnosed with type 1 and 97 with type 2 ovarian cancer; other cases had unknown histotype. Distributions of ovarian cancer risk factors were generally in the expected directions for cases and controls.

**Table 1.**
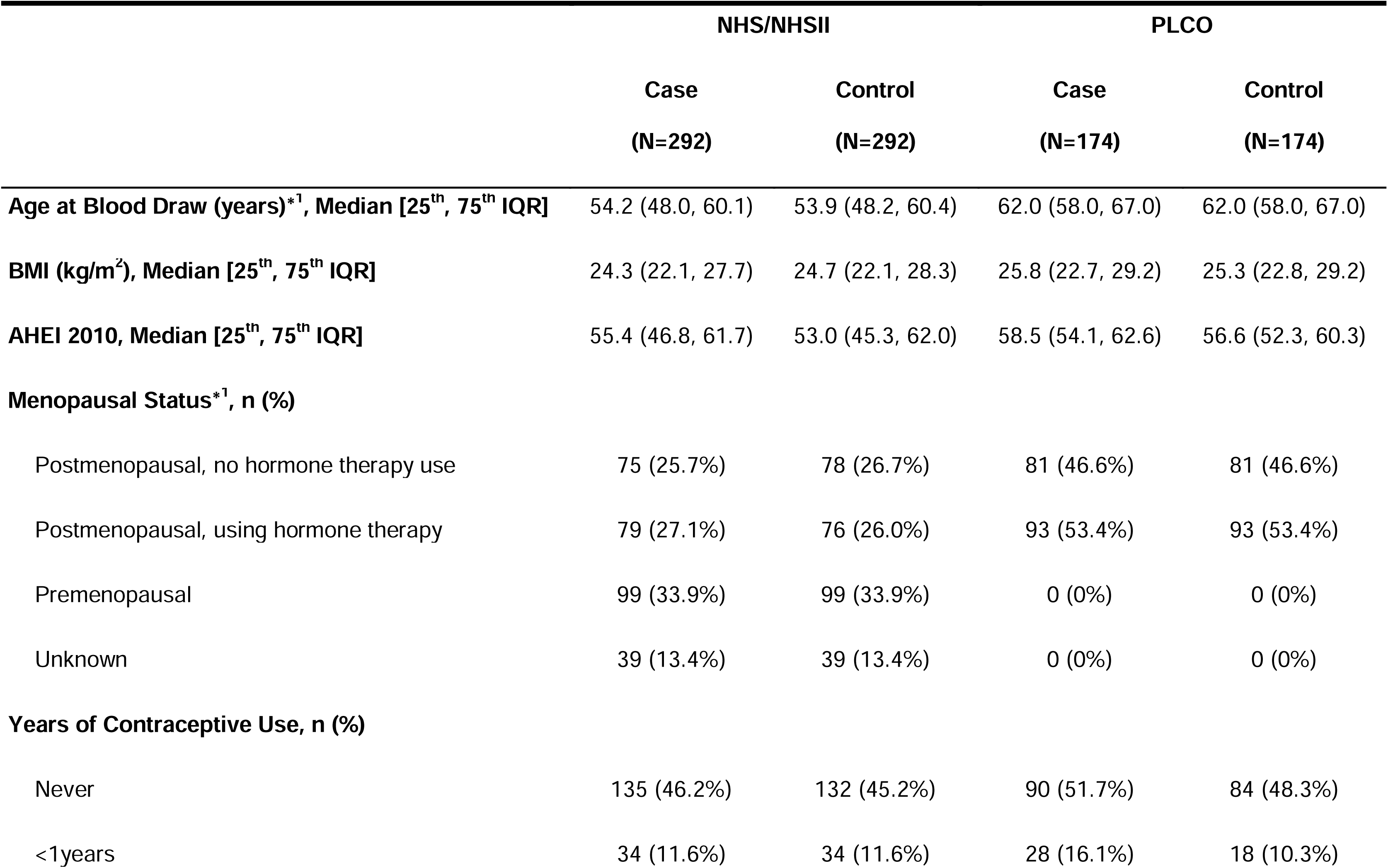
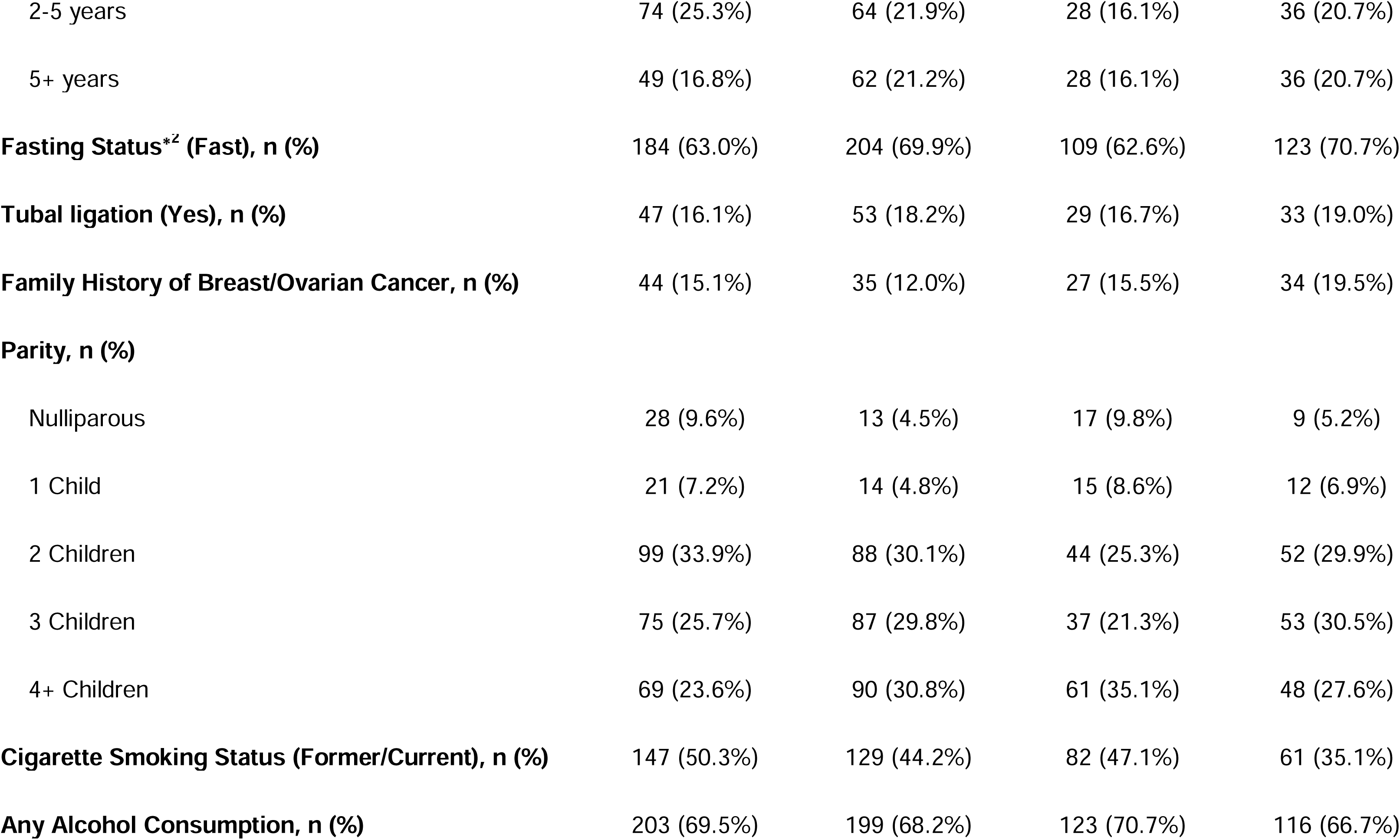
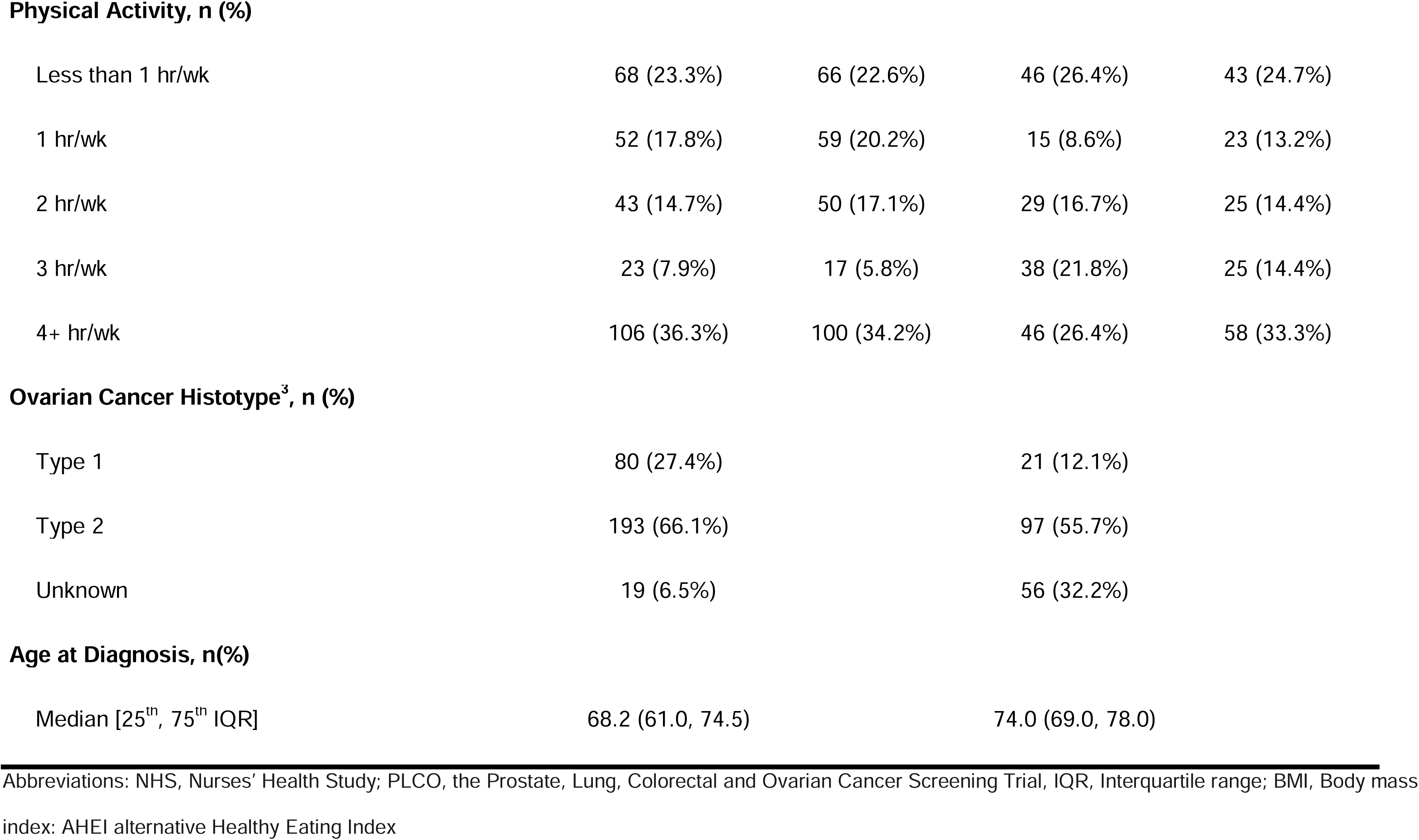
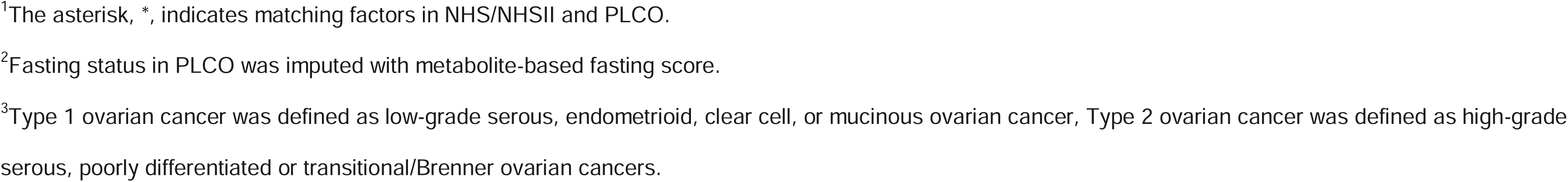
Descriptive characteristics of matched cases and controls nested within NHS/NHSII and PLCO at the time of blood draw.

|  | NHS/NHSII |  | PLCO |  |
| --- | --- | --- | --- | --- |
|  | Case<br>(N=292) | Control<br>(N=292) | Case<br>(N=174) | Control<br>(N=174) |
| **Age at Blood Draw (years)\*<sup>1</sup>, Median [25<sup>th</sup>, 75<sup>th</sup> IQR]** | 54.2 (48.0, 60.1) | 53.9 (48.2, 60.4) | 62.0 (58.0, 67.0) | 62.0 (58.0, 67.0) |
| **BMI (kg/m<sup>2</sup>), Median [25<sup>th</sup>, 75<sup>th</sup> IQR]** | 24.3 (22.1, 27.7) | 24.7 (22.1, 28.3) | 25.8 (22.7, 29.2) | 25.3 (22.8, 29.2) |
| **AHEI 2010, Median [25<sup>th</sup>, 75<sup>th</sup> IQR]** | 55.4 (46.8, 61.7) | 53.0 (45.3, 62.0) | 58.5 (54.1, 62.6) | 56.6 (52.3, 60.3) |
| **Menopausal Status\*<sup>1</sup>, n (%)** |  |  |  |  |
| Postmenopausal, no hormone therapy use | 75 (25.7%) | 78 (26.7%) | 81 (46.6%) | 81 (46.6%) |
| Postmenopausal, using hormone therapy | 79 (27.1%) | 76 (26.0%) | 93 (53.4%) | 93 (53.4%) |
| Premenopausal | 99 (33.9%) | 99 (33.9%) | 0 (0%) | 0 (0%) |
| Unknown | 39 (13.4%) | 39 (13.4%) | 0 (0%) | 0 (0%) |
| **Years of Contraceptive Use, n (%)** |  |  |  |  |
| Never | 135 (46.2%) | 132 (45.2%) | 90 (51.7%) | 84 (48.3%) |
| <1years | 34 (11.6%) | 34 (11.6%) | 28 (16.1%) | 18 (10.3%) |
|  | Case<br>(N=292) | Control<br>(N=292) | Case<br>(N=174) | Control<br>(N=174) |
| 2-5 years | 74 (25.3%) | 64 (21.9%) | 28 (16.1%) | 36 (20.7%) |
| 5+ years | 49 (16.8%) | 62 (21.2%) | 28 (16.1%) | 36 (20.7%) |
| **Fasting Status\*<sup>z</sup> (Fast), n (%)** | 184 (63.0%) | 204 (69.9%) | 109 (62.6%) | 123 (70.7%) |
| **Tubal ligation (Yes), n (%)** | 47 (16.1%) | 53 (18.2%) | 29 (16.7%) | 33 (19.0%) |
| **Family History of Breast/Ovarian Cancer, n (%)** | 44 (15.1%) | 35 (12.0%) | 27 (15.5%) | 34 (19.5%) |
| **Parity, n (%)** |  |  |  |  |
| Nulliparous | 28 (9.6%) | 13 (4.5%) | 17 (9.8%) | 9 (5.2%) |
| 1 Child | 21 (7.2%) | 14 (4.8%) | 15 (8.6%) | 12 (6.9%) |
| 2 Children | 99 (33.9%) | 88 (30.1%) | 44 (25.3%) | 52 (29.9%) |
| 3 Children | 75 (25.7%) | 87 (29.8%) | 37 (21.3%) | 53 (30.5%) |
| 4+ Children | 69 (23.6%) | 90 (30.8%) | 61 (35.1%) | 48 (27.6%) |
| **Cigarette Smoking Status (Former/Current), n (%)** | 147 (50.3%) | 129 (44.2%) | 82 (47.1%) | 61 (35.1%) |
| **Any Alcohol Consumption, n (%)** | 203 (69.5%) | 199 (68.2%) | 123 (70.7%) | 116 (66.7%) |
|  | Case<br>(N=292) | Control<br>(N=292) | Case<br>(N=174) | Control<br>(N=174) |
| **Physical Activity, n (%)** |  |  |  |  |
| Less than 1 hr/wk | 68 (23.3%) | 66 (22.6%) | 46 (26.4%) | 43 (24.7%) |
| 1 hr/wk | 52 (17.8%) | 59 (20.2%) | 15 (8.6%) | 23 (13.2%) |
| 2 hr/wk | 43 (14.7%) | 50 (17.1%) | 29 (16.7%) | 25 (14.4%) |
| 3 hr/wk | 23 (7.9%) | 17 (5.8%) | 38 (21.8%) | 25 (14.4%) |
| 4+ hr/wk | 106 (36.3%) | 100 (34.2%) | 46 (26.4%) | 58 (33.3%) |
| **Ovarian Cancer Histotype<sup>3</sup>, n (%)** |  |  |  |  |
| Type 1 | 80 (27.4%) |  | 21 (12.1%) |  |
| Type 2 | 193 (66.1%) |  | 97 (55.7%) |  |
| Unknown | 19 (6.5%) |  | 56 (32.2%) |  |
| **Age at Diagnosis, n(%)** |  |  |  |  |
| Median [25<sup>th</sup>, 75<sup>th</sup> IQR] | 68.2 (61.0, 74.5) |  | 74.0 (69.0, 78.0) |  |

Abbreviations: NHS, Nurses' Health Study; PLCO, the Prostate, Lung, Colorectal and Ovarian Cancer Screening Trial, IQR, Interquartile range; BMI, Body mass index; AHEI alternative Healthy Eating Index
<sup>1</sup>The asterisk, \*, indicates matching factors in NHS/NHSII and PLCO.
<sup>2</sup>Fasting status in PLCO was imputed with metabolite-based fasting score.
<sup>3</sup>Type 1 ovarian cancer was defined as low-grade serous, endometrioid, clear cell, or mucinous ovarian cancer, Type 2 ovarian cancer was defined as high-grade serous, poorly differentiated or transitional/Brenner ovarian cancers.

The highest MDS quintile, compared to the lowest MDS quintile, was significantly associated with increased ovarian cancer risk across all three models (Model 1: OR=1.53, 95%CI:1.00-2.32, p_trend_=0.09; Model 2: OR=1.58, 95%CI: 1.03-2.42, p_trend_=0.07; Model 3: OR=1.62, 95%CI: 1.03-2.54, p_trend_=0.07) (**Figure 2**, **Supplementary Table 3**). We further examined the association of the 19 MDS metabolites and ovarian cancer risk (**Supplementary Table 4**). We observed that the top versus bottom quintile of circulating levels of pseudouridine (Model 3: OR=1.71, 95%CI: 1.06-2.75, p_trend_=0.12) and N2,N2-dimethylguanosine (Model 3: OR=1.59, 95%CI: 1.01-2.49, p_trend_=0.03) were associated with increased ovarian cancer risk.

**Figure 2.**
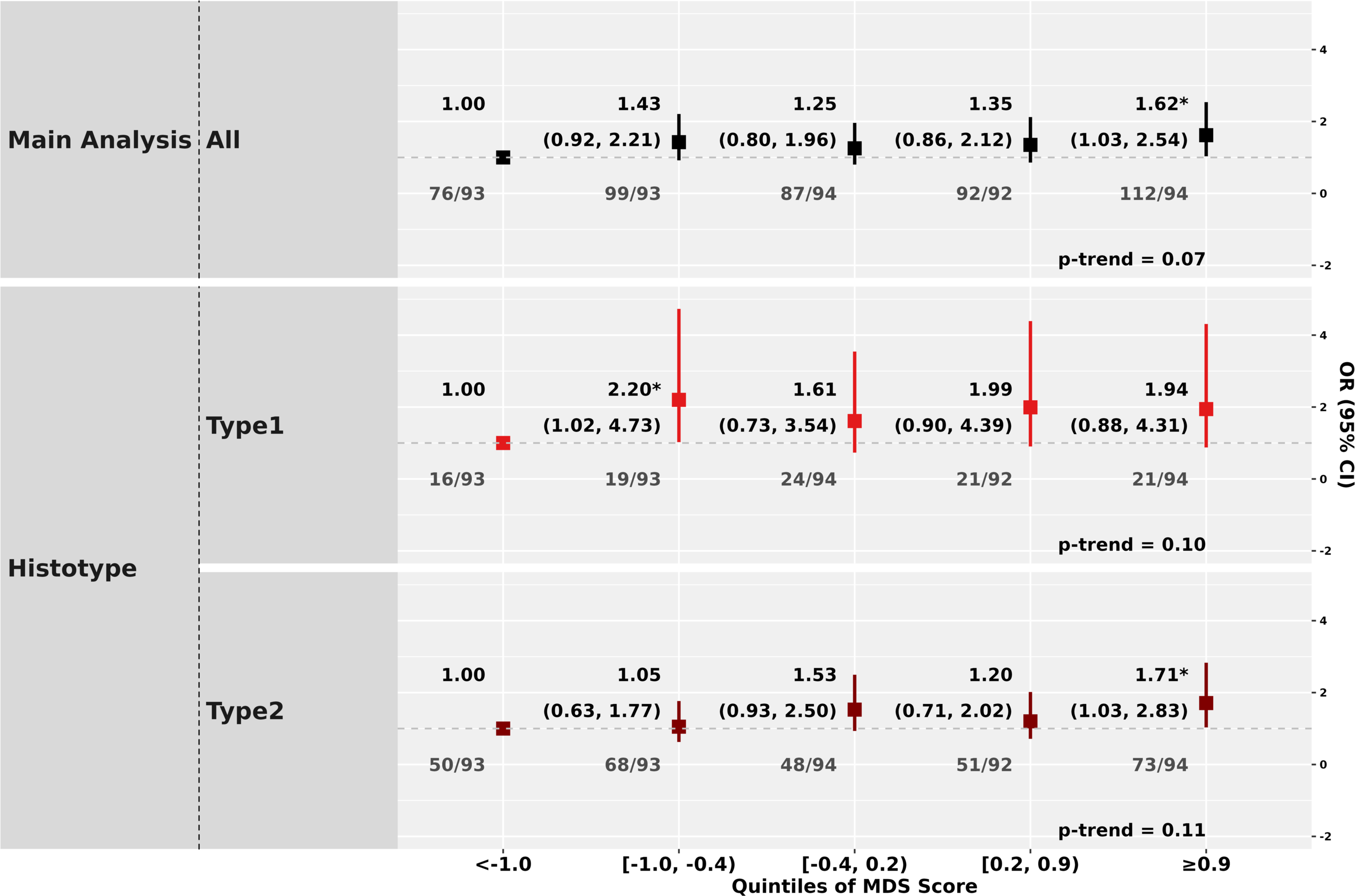
Association of the Metabolite-based Distress Score with Ovarian Cancer Risk Overall and by Histotype. This plot presents the association between quintiles of the metabolite-based distress score (MDS) and ovarian cancer risk evaluated using unconditional logistic regression models, adjusted for matching factors (age at blood collection, study center, fasting status, menopausal status), established ovarian cancer risk factors (race/ethnicity, tubal ligation, oral contraceptive use, parity, family history of breast/ovarian cancer), and lifestyle factors (BMI, smoking status, alcohol consumption, alternative Health Eating Index, physical activity). Odds ratios (ORs) and 95% confidence intervals (CIs) are shown for the main analysis (conducted among ovarian cancer cases with blood collected ≥3 years prior to diagnosis and controls), and across subgroups defined by ovarian cancer histotype (type 1: low-grade serous, endometrioid, clear cell, mucinous; type 2: high-grade serous, poorly differentiated, transitional/Brenner). Each quintile of MDS is compared to the lowest quintile. Samples sizes (cases/controls) are shown below each quintile. Heterogeneity of MDS quintile associations across tumor subtypes was evaluated by likelihood ratio tests comparing a null model in which the association for each MDS quintile was constrained to be equal across case groups to an alternative model in which the MDS quintile associations were allowed to vary across groups based on the trend test. From the second to the last quintiles, the p for heterogeneity is 0.90, 0.07, 0.23, 0.76.

Across ovarian cancer histotypes (**Figure 2**, **Supplementary Table 3**), we only observed statistically significant positive associations for type 2 tumors when comparing the top versus bottom MDS quintile (Model 3: OR=1.71, 95%CI: 1.03-2.83, p_trend_=0.11); however, the test for heterogeneity for type 1 versus type 2 tumors was not statistically significant (p=0.97). The ORs for risk of type 1 tumors ranged from 1.61 (Q3) to 2.20 (Q2) compared to the bottom quintile, with a p_trend_=0.10; however, due to small sample sizes, most estimates were not statistically significant. Among individual MDS metabolites (**Supplementary Table 4**), being in the highest versus lowest quintile of C34:3 PC (Model 3: OR=1.80, 95%CI: 1.01-3.22, p_trend_=0.11) N4-acetylcytidine (Model 3: OR=1.29, 95%CI: 1.04-1.59, p_trend_=0.02), and N2,N2-dimethylguanosine (Model 3: OR=1.25, 95%CI: 1.02-1.54, p_trend_=0.03) was associated with increased type 2 ovarian cancer risk. Additionally, the highest versus lowest quintile of C16:0 ceramide was associated with increased type 1 ovarian cancer risk with a significant linear trend (Model 3: OR=5.07, 95%CI: 1.31-19.59, p_trend_=0.03).

Among premenopausal women, an elevated ovarian cancer risk was observed for the 3rd to 4th versus the lowest MDS quintile (Model 3: 3rd-quintile: OR=3.00, 95%CI: 1.05- 8.54; 4th-quintile: OR=3.00, 95%CI: 1.08-8.31; p_trend_=0.08), with a suggestive association for the highest quintile (Model 3: OR=2.13, 95%CI: 0.72-6.08) (**Figure 3**, **Supplementary Table 3**). No clear associations were observed for postmenopausal women (Model 3, top versus bottom quintile: OR=1.47, 95%CI: 0.85-2.54). There was no suggestion of heterogeneity in top versus bottom quintile by menopausal status (p=0.29). For individual MDS metabolites (**Supplementary table 4**), the 2nd, 3rd, and 4th quintile of 3-methylxanthine (Model 3: 2nd-quintile: OR=3.67, 95%CI: 1.19-11.30; 3rd-quintile: OR=3.08, 95%CI: 1.03-9.22; 4th-quintile: OR=3.05, 95%CI: 1.01-9.19) were associated with increased risk of ovarian cancer among premenopausal women, with a suggestive association for the highest quintile (OR=2.75, 95%CI: 0.90-8.35; p_trend_=0.17).

**Figure 3.**
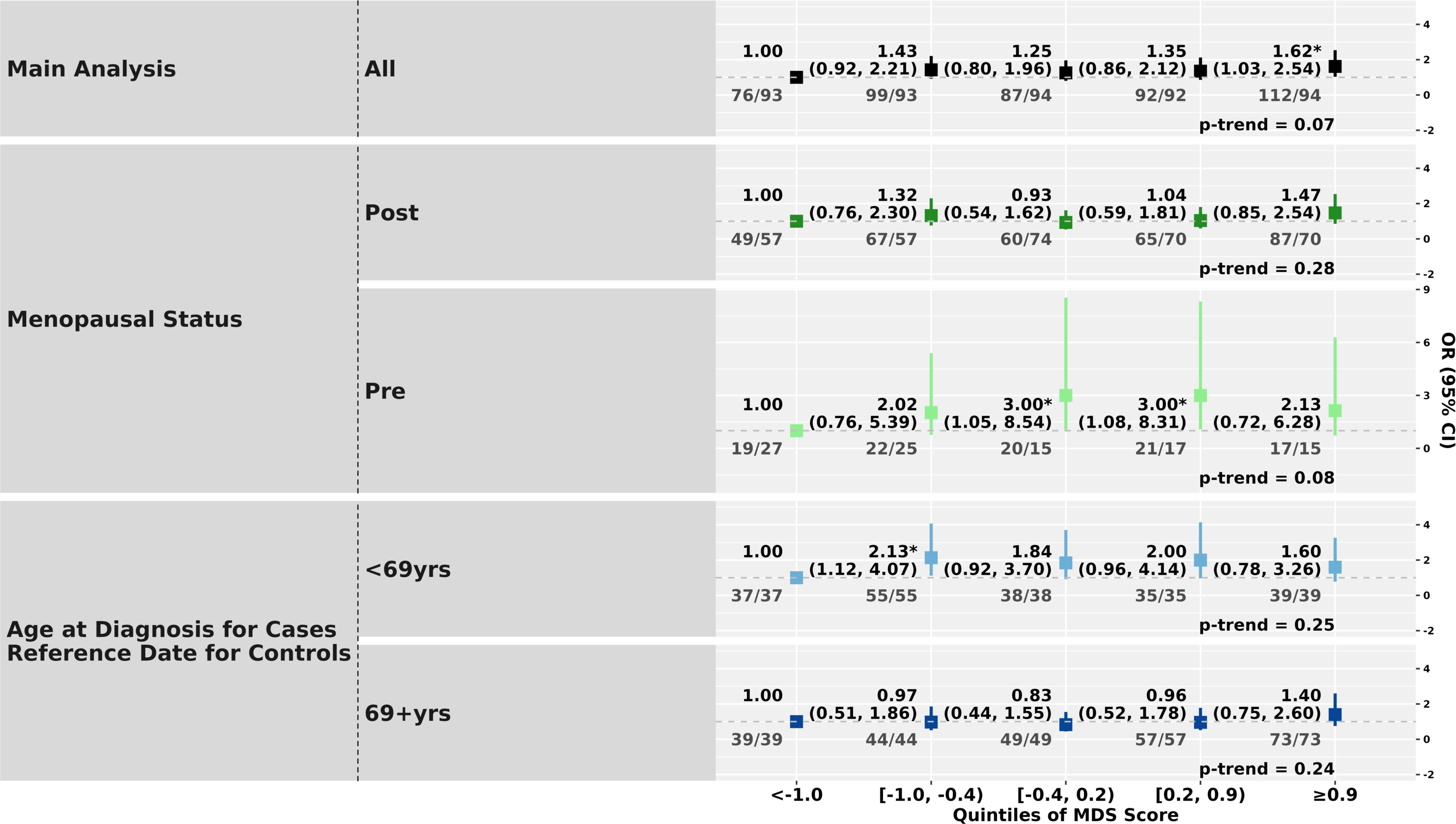
Associations of Metabolite-based Distress Score and Ovarian Cancer Risk Stratified by Patient Characteristics. This plot presents the association between quintiles of the metabolite-based distress score and ovarian cancer risk evaluated using unconditional logistic regression models, adjusted for matching factors (age at blood collection, study center, fasting status, and menopausal status); established ovarian cancer risk factors (race/ethnicity, tubal ligation, oral contraceptive use, parity, and family history of breast/ovarian cancer); and lifestyle factors (BMI, smoking status, alcohol consumption, dietary factors, and physical activity). Odds ratios (ORs) and 95% confidence intervals (CIs) are shown for the main analysis (conducted among ovarian cancer cases with blood collected ≥3 years prior to diagnosis and controls), and across subgroups defined by menopausal status (premenopausal, postmenopausal) and age at diagnosis (<69 years, ≥69 years). Each quintile of MDS is compared to the lowest quintile. Samples sizes (cases/controls) are shown below each quintile. Heterogeneity of MDS quintiles in associations across tumor subtypes were evaluated by likelihood ratio tests comparing a null model in which the associations for each MDS quintile were constrained to be equal across case groups, and an alternative model in which the MDS quintile associations were allowed to vary across groups based on the trend test. From the second to the last quintiles, the p for heterogeneity is 0.46, 0.05, 0.07, 0.55 for menopausal status and 0.09, 0.10, 0.13 and 0.78 for age at diagnosis.

Among women diagnosed at less than the median age at diagnosis (69 years old), there was a suggestion of an elevated ovarian cancer risk across quintiles 2-5 (Model 3: ORs=2.13, 1.84, 2.00, 1.60, respectively) (**Figure 3**, **Supplementary Table 3**). The association in the top versus bottom quintile for those diagnosed at ≥69 years was 1.40 (95%CI: 0.75-2.60; p-heterogeneity=0.91). For individual MDS metabolites (**Supplementary Table 4**), the 5th versus 1st quintile of C34:3 PC (Model 3: OR=2.31, 95%CI: 1.08-4.94, p_trend_=0.20) was significantly associated with increased ovarian cancer risk and a significant positive linear trend was observed for N4-acetylcytidine (p_trend_=0.01) in the younger age group.

Among participants who had blood collected more than 12 years before diagnosis, the top versus bottom MDS quintile was associated with increased risk of ovarian tumors (Model 3: OR=1.90, 95%CI: 1.10-3.28, p_trend_=0.02), but not among those had blood collected 3-12 years before diagnosis (Model 3: OR=1.36, 95%CI: 0.78-2.37, p_trend_=0.56; p-heterogeneity=0.30) (**Supplementary Figure 3, Supplementary Table 3**). For individual MDS metabolites (**Supplementary Table 4**), significant positive linear trends were observed for pseudouridine (Model 3: OR=2.36, 95%CI: 1.16-4.77, p_trend_=0.03) and N4-acetylcytidine (p_trend_=0.01) among those had blood collected 3-12 years before diagnosis. No differences were observed when we stratified by fasting status (**Supplementary Tables 3 and 4**).

## Discussion

We conducted the first multi-cohort analysis to investigate the association between a pre-diagnostic, metabolite-based distress score and ovarian cancer risk. Overall, we observed that women in the highest quintile of the MDS had an increased risk of ovarian cancer compared to those in the lowest quintile, with a non-significant trend, suggesting a threshold effect. While we only observed significant associations for type 2 tumors, there was no significant heterogeneity by histotype and power was limited for the type 1 tumors, despite having 3 cohorts. Although based on relatively small numbers, the associations may be stronger among premenopausal women at blood collection and among those diagnosed before age 69, with a suggestive increased risk from quartiles 2-5 compared to the first quartile. The observed associations appeared to be largely driven by positive associations with several individual MDS metabolites, including pseudouridine and N2,N2-dimethylguanosine.

We observed a positive association between MDS and ovarian cancer risk, whereby women in the highest quintile, representing the highest levels of distress as reflected by circulating metabolites, were at increased ovarian cancer risk. The associations were not significant in quintiles 2-4, with ORs ranging from 1.25-1.43, suggesting the possibility of a threshold effect. Our observation that chronic psychological stress is associated with ovarian cancer risk aligns with prior epidemiologic findings. One study reported socially isolated and widowed women had an increased risk of ovarian cancer.^12^ Another study found that women with three or more distress-related psychosocial factors, including depression, anxiety, and post-traumatic stress disorder (PTSD), had higher ovarian cancer risk.^27^ When focusing on PTSD, women experiencing six or more PTSD symptoms showed an elevated risk of ovarian cancer.^11^ Collectively, these studies reinforce chronic psychological stress as a potential risk factor for ovarian cancer and suggest that more extreme psychological distress is associated with the highest risk.

When examining individual MDS metabolites, pseudouridine and N2,N2- dimethylguanosine drove the association of MDS and ovarian cancer risk. A positive association between circulating pseudouridine levels and ovarian cancer risk has been reported previously, including in a study included some samples from NHS/NHSII.^19,28,29^ Pseudouridine is involved in RNA stability and regulation.^30^ Elevated pseudouridine was reported in individuals with major depressive disorder, likely reflecting upregulated pseudouridylation driven by overexpressed small nucleolar RNAs (snoRNAs).^31^ Specifically, the snoRNA-induced pseudouridylation is reported to promote ovarian cancer progression via cancer-associated alternative splicing.^32^ Together, these findings suggest psychological distress may increase ovarian cancer risk through upregulation of pseudouridylation pathways. N2,N2-dimethylguanosine, a methylated nucleoside that forms during the degradation of transfer RNA, is essential for RNA folding and stability.^33^ Elevated plasma levels of N2,N2-dimethylguanosine was associated with colorectal cancer risk in men,^34^ likely reflecting increased cellular metabolism and accelerated RNA turnover in tumor cells.^35,36^ Our findings suggest that psychological distress may drive metabolomic alterations that disrupt RNA processing and stability, ultimately contributing to cancer development.

When stratifying by ovarian cancer histotypes, we observed an association of MDS with ovarian cancer risk in the more common type 2 tumors. This finding is consistent with prior epidemiologic studies reporting that self-reported depression was associated with increased risk of type 2 tumors.^37^ One proposed mechanism is that chronic psychological distress activates the sympathetic nervous system, leading to elevated levels of norepinephrine. Experimental studies have shown that norepinephrine can promote carcinogenic processes in fallopian tube precursor-like cells with p53 mutations, a predominate somatic mutation of type 2 tumors.^37–39^ These findings suggest that type 2 tumors may be biologically susceptible to the effects of stress-related hormonal signaling. That said, the power was limited for analyses stratified by tumor type. Additional research is needed to increase the sample size of type 1 tumors to more precisely evaluate the association, especially since studies of self-reported distress have observed associations with both type 1 and type 2 tumors.

When examining the individual MDS metabolites, C34:3 PC, N4-acetylcytidine and N2,N2-dimethylguanosine were associated with risk of type 2 tumors. A recent prospective cohort study identified a positive association between circulating C34:3 PC levels and PTSD.^40^ Additionally, elevated C34:3 PC levels are associated with increased ovarian cancer risk^41^. These findings suggest that chronic distress may dysregulate C34:3 PC metabolism, leading to inflammation^40^ leading to higher ovarian cancer risk.^11^ Further, N4-acetylcytidine, a post-transcriptional RNA modification, envolved in mRNA stability, splicing, and translation efficiency.^42^ N-acetyltransferase 10 (NAT10), which catalyzes N4-acetylcytidine, is associated with brain-related functions in animal studies, including memory^43^ and depression.^44^ Notably, studies have shown that overexpression of NAT10 in the hippocampus leads to enhanced depressive and anxiety-like behaviors in mice^44^ suggesting that overexpression of NAT10 may be associated with depression through upregulating N4-acetylcytidine levels. Moreover, N4-acetylcytidine is associated with tumor development. Elevated N4-acetylcytidine levels, mediated by NAT10, upregulate the expression of oncogenes, CAPRIN1 and ACOT7, in serous (i.e., type 2) ovarian tumors.^45,46^ Our findings are consistent with prior evidence that N4- acetylcytidine is associated with both depression and ovarian cancer, supporting a pathway in which chronic phycological distress promotes the activation of downstream oncogenic processes.

Interestingly, C16:0 ceramide showed a positive association with risk of type 1 tumors. Prior studies demonstrated that chronic stress activates acid sphingomyelinase, elevating hepatic C16:0 ceramide levels in mice.^47^ A prospective cohort study, that included a subset of samples in this study, also reported a suggestive positive association between circulating C16:0 ceramide and type 1 ovarian cancer risk,^20^ which is consistent with a Mendelian randomization analysis showing a positive association of ceramides with ovarian cancer risk.^48^ Notably, C16-ceramide is synthesized by Ceramide synthase 5, which activates β-catenin and SOAT1 and promotes tumor cell proliferation.^49^ Our results reinforce these findings and suggest that ceramide accumulation may be a distress-related metabolic signature linked to type 1 ovarian carcinogenesis.

In stratified analyses, we observed a stronger association of MDS with ovarian cancer risk among premenopausal women and women <69 years at diagnosis, although sample sizes were small and there was no significant heterogeneity by menopausal status or age at diagnosis. These findings are consistent with previous research suggesting slightly stronger associations of depression and PTSD with increased ovarian cancer risk in premenopausal women,^9,11^ although anxiety was more strongly associated with risk in postmenopausal women.^10^ This could be due to distinct biological mechanisms during reproductive years. For instance, post-ovulatory wound healing, a pathway implicated in ovarian cancer development, is only active during reproductive age.^50^ Notably, evidence suggests that norepinephrine in the peritoneal cavity increases during ovulation and that norepinephrine exposure of preneoplastic lesions led to increased cell viability and spheroid formation, as well as resistance to anoikis.^38,39^ Additional work should also evaluate the potential for hormonal interactions, given the vastly different estrogenic environment by menopausal status.

When examining the individual MDS metabolites, 3-methylxanthine, a purine derivative in caffeine metabolism, was associated with ovarian cancer risk among premenopausal women. While direct studies examining the association between 3-methylxanthine and psychological distress are limited, several investigations have explored links between coffee consumption and distress-related outcomes, with mixed findings. For instance, moderate coffee intake has been associated with reduced depressive symptoms^51^ and lower suicide risk,^51,52^ whereas, in rare cases, high doses of caffeine have been shown to induce anxiety.^51^ Additionally, association between 3-methylxanthine and psychological distress may extend beyond caffeine intake. A prior epidemiological study reported a positive relationship between circulating 3-methylxanthine levels and distress even after adjusting for dietary caffeine intake.^16^ Conversely, a mouse model of ovarian cancer specifically showed that 3-methylxanthine enhanced apoptosis with cisplatin treatment via the dopamine receptor pathways.^53^ 3-methylxanthine’s potential role in ovarian cancer development, particularly among premenopausal women, warrants further investigation.

This study has several notable strengths. It represents the largest multi-cohort investigation to date examining the association between depression-related metabolomic dysregulation and ovarian cancer risk. The cohort studies included long-term follow-up and comprehensive covariate data, enhancing the robustness of the analysis. However, several limitations should be acknowledged. First, both cohorts measured 19 of the 20 metabolites included in the original MDS. Although efforts were made to recover the missing MDS metabolite from the pool of unidentified compounds, this may have attenuated the observed association between MDS and ovarian cancer risk. Second, despite including 932 participants in the analysis, the study power was limited in the stratified analyses.

We identified a significant association between a pre-diagnostic, metabolite-based distress score and ovarian cancer risk. Using a biomarker-based measure of distress has advantages in overcoming potential issues with self-reported distress, such as social desirability in reporting, as well as better reflecting inter-individual variability in the biologic effects of distress. The MDS provides a measurement of biologic dose that may be less impacted by confounding factors. The MDS-ovarian cancer relationship generally was robust across different tumor types and across varying subgroups, although there was some evidence greater susceptibility to distress-related metabolic dysfunction regarding ovarian cancer development in younger women. Further, several individual MDS metabolites, pseudouridine and N2,N2-dimethylguanosine were significantly associated with ovarian cancer risk, implicating pathways related to RNA modification. Together, these findings support the hypothesis that chronic psychological distress may influence ovarian cancer development through metabolomic alterations, warranting further research to elucidate underlying biological mechanisms and the populations most impacted by the carcinogenic effects of distress.

## Data availability statement

Due to participant confidentiality and privacy concerns, data cannot be shared publicly and requests to access NHS/NHSII data must be submitted in writing. According to standard controlled access procedures, applications to use NHS/NHSII resources will be reviewed by our External Collaborations Committee to verify that the proposed use maintains the protection of the privacy of participants and the confidentiality of the data. Investigators wishing to use NHS/NHSII data are asked to submit a brief description of the proposed project. Please see https://www.nurseshealthstudy.org/researchers (contact) for details.

## Acknowledgements

This study was supported by the DoD RPPR grant HT9425-23-1- 0236. The Nurses’ Health Studies infrastructure was supported by UM1CA186107, P01CA87969, R01CA49449, U01CA176726, R01CA67262. The authors thank the National Cancer Institute for providing the human material collected by the Prostate, Lung, Colorectal and Ovarian Cancer (PLCO) Screening Trial.

The authors would like to acknowledge the contribution to this study from central cancer registries supported through the Centers for Disease Control and Prevention’s National Program of Cancer Registries (NPCR) and/or the National Cancer Institute’s Surveillance, Epidemiology, and End Results (SEER) Program. Central registries may also be supported by state agencies, universities, and cancer centers. Participating central cancer registries include the following: Alabama, Alaska, Arizona, Arkansas, California, Delaware, Colorado, Connecticut, Florida, Georgia, Hawaii, Idaho, Indiana, Iowa, Kentucky, Louisiana, Maine, Maryland, Massachusetts, Michigan, Mississippi, Montana, Nebraska, Nevada, New Hampshire, New Jersey, New Mexico, New York, North Carolina, North Dakota, Ohio, Oklahoma, Oregon, Pennsylvania, Puerto Rico, Rhode Island, Seattle SEER Registry, South Carolina, Tennessee, Texas, Utah, Virginia, West Virginia, Wyoming.

## Author contributions

Conceptualization, methodology: N.L., S.S.T., O.A.Z. Data analysis: N.L. Study design, supervision: S.S.T., O.A.Z. Writing-original draft: N.L. Writing- review and editing: N.L., R.J., G.M., A.H.E., B.T., J.A., M.K.T., K.L.T., C.B.C., S.S.T., O.A.Z.

