## Supplementary Materials for "The Role of Distress-related Metabolic Dysfunction in Ovarian Cancer Development: a pooled case-control study"

### ***Metabolomics assay***

Metabolomic data was measured using complimentary liquid chromatography tandem mass spectrometry (LC-MS) methods designed to measure polar metabolites and lipids as well as free fatty acids on four platforms, including polar and non-polar lipids (C8-pos), and cationic polar metabolites (HILIC-pos). The metabolomics platforms measure metabolites with different features of ionization, polarity, and hydrophilicity. Hydrophilic interaction liquid chromatography (HILIC) analyses of water-soluble metabolites in the positive ionization mode (HILIC-pos) were conducted using an LC-MS system comprised of a Shimadzu Nexera X2 U-HPLC (Shimadzu Corp.; Marlborough, MA) coupled to a Q Exactive mass spectrometer (Thermo Fisher Scientific; Waltham, MA). Plasma lipids (C8-pos) were profiled using a Shimadzu Nexera X2 U-HPLC (Shimadzu Corp.; Marlborough, MA). Metabolites of intermediate polarity (C18-neg), including free fatty acids and bile acids, were profiled using a Nexera X2 U-HPLC (Shimadzu Corp.; Marlborough, MA) coupled to a Q Exactive (Thermo Fisher Scientific; Waltham, MA). Raw data from orbitrap mass spectrometers was processed using TraceFinder 3.3 software (Thermo Fisher Scientific; Waltham, MA) and Progenesis QI (Nonlinear Dynamics; Newcastle upon Tyne, UK) and targeted data from the QTRAP 5500 system were processed using MultiQuant (version 2.1, SCIEX; Framingham, MA). For each method, metabolite identities were confirmed using authentic reference standards or reference samples.

Metabolomics data were collected in two batches. Batch 1 included 691 NHS/NHSII samples, 122 quality control samples, and 10 drift samples and batch 2 included 82 NHS/NHSII samples, 526 PLCO samples, 48 QC samples, and the same 10 drift samples included in batch 1. We performed quality control checks to remove samples that were not collected with robust collection techniques (e.g., processed >24 hours after collection). We excluded metabolites that were redundantly measured across liquid chromatography / mass spectrometry platforms within a batch and retained only one measurement per metabolite (N=48 in batch 1, N=40 in batch 2), that did not meet reproducibility standards (i.e., high coefficient of variation; N=48 in batch 1, N=48 in batch 2), or represented ambiguous isoforms (N=16 in batch 1, N=21 in batch 2). After quality control measures, our dataset contained 477 and 515 metabolites from batches 1 and 2, respectively. Among the 314 metabolites shared between batches, over 90% demonstrated mean coefficients of variation (CVs) below 25% in both, indicating strong technical reproducibility. All MDS metabolites included in this study had CV <25%.

### ***Assessment of covariates***

For the NHS and NHSII cohorts, age, weight, menopausal status, and use of hormone therapy were assessed at the time of blood collection. Height, duration of oral contraceptive use, parity, tubal ligation, and family history of breast or ovarian cancer were obtained from the biennial questionnaires completed prior to blood collection; height and weight were used to calculate body mass index (BMI) at blood collection. For the PLCO cohort, age, BMI, menopausal status and hormone therapy use, duration of oral contraceptive use, parity, tubal ligation, and family history of breast or ovarian cancer were collected at the baseline screening visit.

Physical activity was assessed in both studies. In PLCO, participants reported the number of hours of exercise per week at the baseline screening visit. In NHS/NHSII, participants reported average weekly time spent in eight specific activities, including walking, jogging, running, bicycling, swimming, playing tennis, playing squash or racquetball, and doing calisthenics/aerobics/rowing. Total weekly activity time was summed and categorized into <1, 1-2, 2-3, 3-4, or ≥4 hours per week to match the PLCO categories. In NHS/NHSII, diet was evaluated using the Alternative Healthy Eating Index (aHEI), derived from a food frequency questionnaire assessing average dietary intake over the preceding 12 months.^25^ In PLCO, we calculated aHEI scores using the food frequency questionnaire and a validated method described elsewhere.^26^

### ***Calibration of multi-batch metabolomic data***

To normalize data between batches, we recalibrated batch 2 metabolite values using linear regression and the 10 drift samples measured in both batches, with values in batch 2 regressed on the measured levels from batch 1. Specifically, natural log-transformed metabolite levels in batch 2 were adjusted using regression intercept and beta coefficients for each metabolite. A principal component analysis (**Supplementary** **Figure 1**) was performed to evaluate the effectiveness of recalibration and to confirm improved alignment across batches.

### ***Metabolite-based distress score (MDS) and recalibration of MDS coefficients***

The metabolite-based distress score (MDS) was previously developed by Dr. Balasubramanian in a case-control dataset nested within the Nurses’ Health Study^1^. The dataset consisted of 279 women with prevalent chronic distress (characterized by recurring experiences of high levels of depression and anxiety) and 279 matched controls. A total of 20 metabolites were selected by elastic net as the metabolite-based distress score (MDS). The MDS was created using coefficients from a logistic regression model including the 20 metabolites (using a z-score for normalization) with chronic distress as the outcome. Of the 20 MDS metabolites, 19 and 18 metabolites were measured in batch 1 and batch 2 in our study, respectively. We utilized PAIREDUP-MS^2^, an imputation-based approach that leverages mass-to-charge ratios, and rescued 1 metabolite (C34:3 PC) from the unknown metabolites in batch 2. To optimize the MDS score, we recalculated the regression coefficients using the 19 metabolites, z-score transformed, found in our dataset using the original case-control dataset used in the MDS development. Due to the non-normal distribution of many of the metabolites, our final MDS score was based on a regression model of the 19 metabolites after probit transformation (a non-parametric ranking approach that ensures a normal distribution) on chronic distress. The comparison of the coefficients is presented in **Supplementary Figure 2**. We then calculated MDS for all available samples in the two nested case control studies in NHS/NHSII and PLCO.

### ***Creation of a fasting score to predict fasting status in PLCO.***

In PLCO, no information was collected on fasting status, which is a pre-analytic factor that impacts many metabolites. Thus, we developed a metabolite-based fasting score (MFS) to predict fasting status. A total of 150 metabolites from platforms of polar and non-polar lipids (C8-positive) and cationic polar metabolites (HILIC-positive) from 6,299 postmenopausal healthy controls (to mimic PLCO eligibility criteria) in NHS/NHSII (5,960 from NHS and 339 from NHSII) were included in MFS development. The length of time since the participant last ate was collected in the NHS/NHSII blood questionnaire and collapsed into an ordinal variable (1: <2 hours, 2: 2-4 hours, 3: 5-7 hours, 4: 8-11 hours, and 5: >=12 hours). The NHS/NHSII dataset (n = 6,299) was split in a 70:30 fashion, where 70% of the dataset was sampled to serve as a training set and 30% of the dataset remained as testing set 1. We also created a second testing set with postmenopausal ovarian cancer cases and controls from the ovarian cancer case-control study nested within NHS/NHSII. Elastic net regression was used to select metabolites related to time since last meal in the training set with a 10-fold cross-validation framework, which was then applied to testing sets 1 and 2. The MFS was calculated as the weighted sum of the selected metabolites and participant characteristics with weights equal to the beta coefficients of the selected metabolites, age at blood draw and body mass index (BMI) from a linear regression model regressed on time since last meal. Optimal cut-off of predicted time since last meal was calculated to maximize sensitivity and specificity in testing set 1, where the optimal cut-off for predicted fasting status was a time since last meal of >8 hours, matching our definition of fasting in the training set. Summary statistics of model development are presented in **Supplementary Table 1** and the developed fasting score is presented in **Supplementary Table 2**.

**Supplementary Figure 1. Principal component analysis of recalibrated metabolomic datasets.** Scatter plot showing the first two principal components (PC1 and PC2) derived from metabolomics data across three groups: Batch 1 NHS (green), Batch 2 NHS (orange), and Batch 2 PLCO (purple). Each point represents an individual sample. Left plot is pre-calibration, and right plot is post-calibration. The distribution of samples in the PCA space reflects overall similarities in global metabolomics profiles.

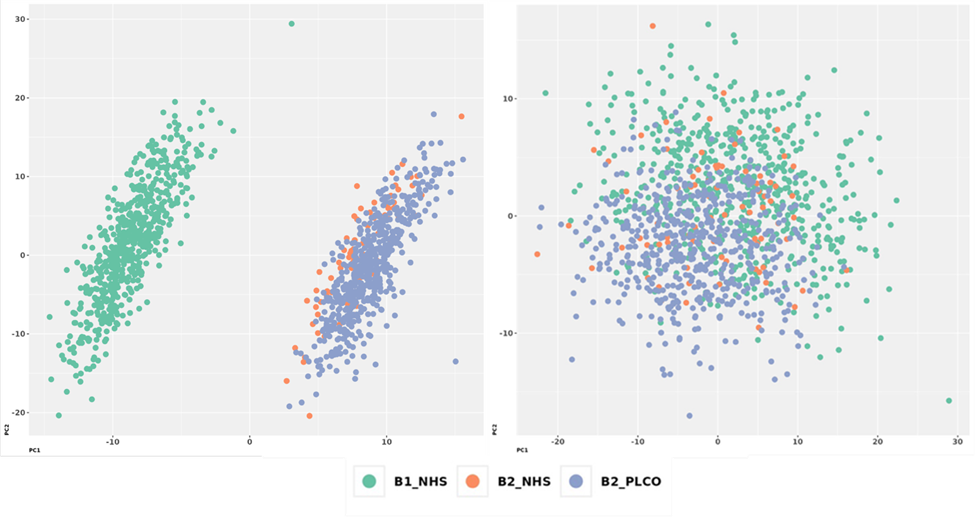

**Supplementary Figure 2.** **Comparison of original and recalibrated MDS score coefficients.** This heatmap displays the standardized beta coefficients of individual metabolites included in three versions of the Metabolite Distress Score: MDS 19 (19 metabolites measured in NHS with metabolite values normalized by z-score of log-transformed metabolite values), MDS 19 probit (19 metabolites measured in NHS with metabolite values normalized by probit score), and MDS 20 (the original model with metabolite values normalized by z-score of log-transformed metabolite values). Each cell represents the coefficient of each metabolite in the corresponding model. Positive associations are shown in red, and negative associations are shown in blue, with intensity reflecting the magnitude of the beta coefficient.

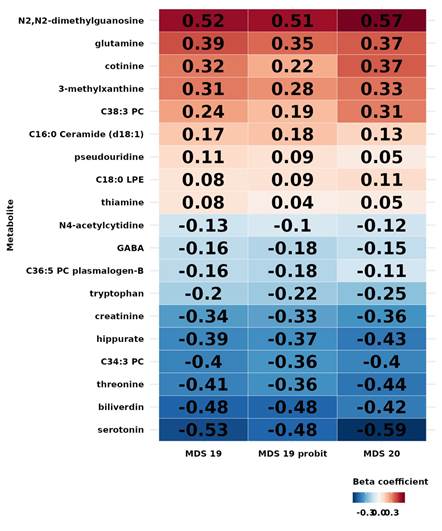

**Supplementary Figure 3. Associations of Metabolite-based Distress Score and Ovarian Cancer Risk Stratified by Time between Blood Draw and Diagnosis**

This plot presents the association between quintiles of the metabolite-based distress score and ovarian cancer risk evaluated using unconditional logistic regression models, adjusted for matching factors (age at blood collection, study center, fasting status, and menopausal status); established ovarian cancer risk factors (race/ethnicity, tubal ligation, oral contraceptive use, parity, and family history of breast/ovarian cancer); and lifestyle factors (BMI, smoking status, alcohol consumption, dietary factors, and physical activity). Odds ratios (ORs) and 95% confidence intervals (CIs) are shown for the main analysis (conducted among ovarian cancer cases with blood collected ≥3 years prior to diagnosis and controls), and across subgroups defined by time between blood draw and diagnosis (3-12 years, and 12+ years). Each quintile of MDS is compared to the lowest quintile. Samples sizes (cases/controls) are shown below each quintile.

Heterogeneity of MDS quintiles in associations across tumor subtypes were evaluated by likelihood ratio tests comparing a null model in which the associations for each MDS quintile were constrained to be equal across case groups, and an alternative model in which the MDS quintile associations were allowed to vary across groups based on the trend test. From the second to the last quintiles, the p for heterogeneity is 0.88, 0.81, 0.22, 0.30.

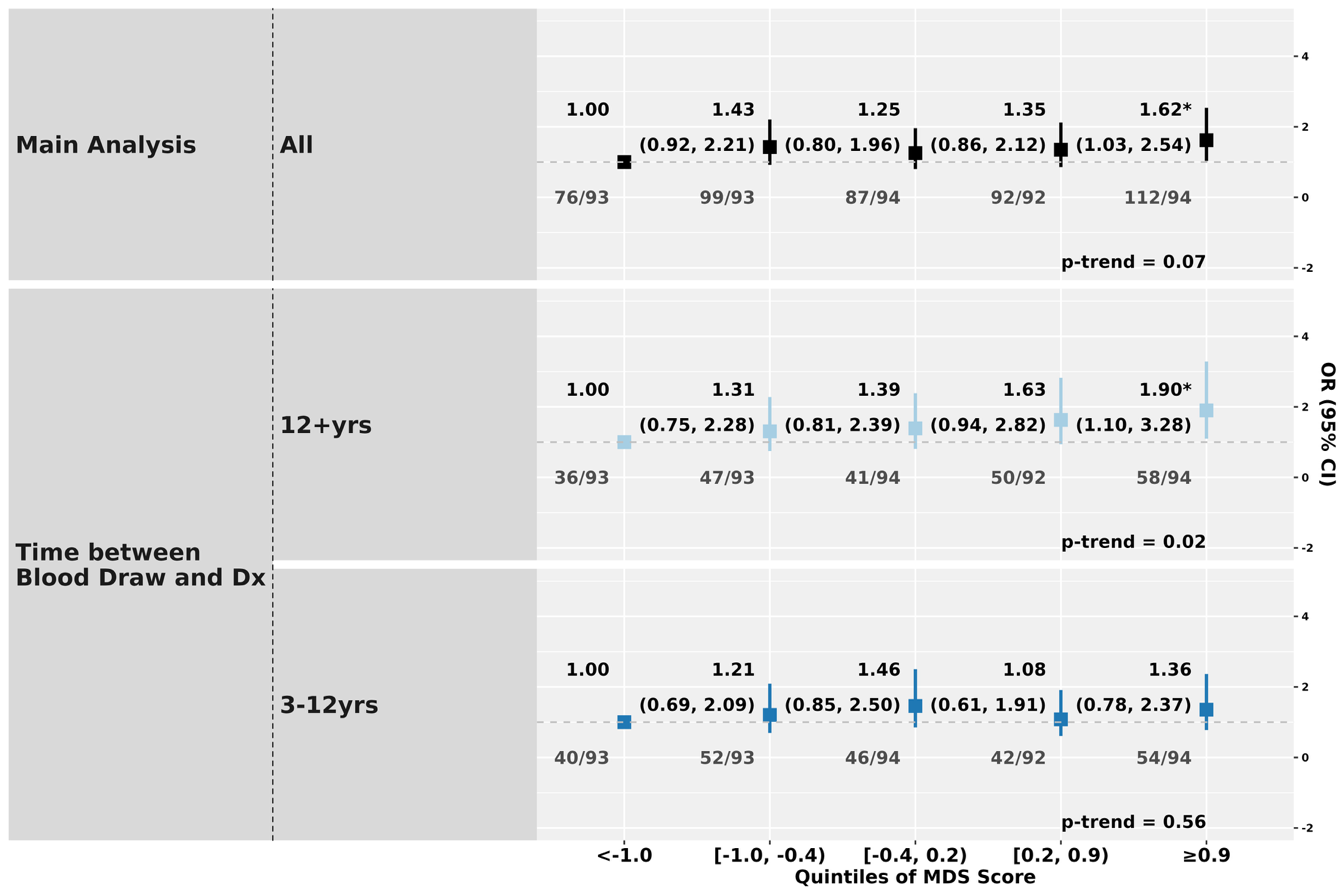
